# Prevalence and causes of blindness and visual impairment in Sudur paschim Province of Nepal

**DOI:** 10.1101/2022.11.24.22282704

**Authors:** Suresh Raj Pant, Ramesh Chandra Bhatta, Subash Bhatta, YuddhaDhoj Sapkota

**Author notes:** **Address for Correspondence:** Suresh Raj Pant, Geta Eye Hospital, Dhangadhi, Nepal. **Authors Contribution**. Data analysis, Literature Review, Manuscript writing (Suresh Raj Pant, Ramesh Chandra Bhatta). Review of the final draft and editing reports (Subash Bhatta, YuddhaDhoj Sapkota). **Coauthors:** Ramesh Chandra Bhatta, Subash Bhatta, Yuddha Dhoj Sapkota.

## Abstract

**Aim:** To determine the prevalence and causes of blindness and visual impairment among aged 50 years and older population in Sudurpaschim Province of Nepal.

**Methods:** A cross sectional study, among population aged 50 years and older using a random multistage cluster sampling procedure. Eligible study participants were enrolled by door-to-door enumeration. Survey team lead by an Ophthalmologist assessed visual acuity, examination of eye and data collection was carried out on tablets installed with mRAAB 7 software.

**Results:** The survey examined 4573 out of 4615 subjects enumerated, with a response rate 99.1%. Out of 4573 subjects, 1995 (43.63%) were male and 2578 (56.37%) were female. Among the examined, 2214 (48.4%) were between 50-59 years. The age- and sex-adjusted prevalence of blindness was 0.6% (95% CI 0.3 – 0.87).The age- and sex-adjusted prevalence of severe visual impairment, moderate visual impairment, and early visual impairment were 1.0% (95% CI 0.6 – 1.3), 5.3% (95% CI 4.5 – 6.2) and 7.0% (95% CI 6.0 – 8.0) respectively. The main causes of bilateral blindness were untreated cataract (36.4%), non-trachomatous corneal opacity (24.2%), glaucoma (21.2%), globe or central nervous system abnormalities (9.1%), age related macular degeneration (6.1%) and other posterior segment diseases (3.0%).

**Conclusion:** Prevalence of blindness was reduced in the province in comparison to the previous survey. Cataract remains the major cause of blindness, severe visual impairment, and moderate visual impairment followed by corneal opacities and glaucoma. The study suggests more attention to be given for cataract, corneal blindness and glaucoma in future interventions to eliminate blindness.

## Introduction

According to World Health Organization (WHO), visual impairment (VI) is an important health problem in both developed and developing countries. It is estimated that 39 million people are blind globally and 246 million have low vision. About 65% of the visually impaired and 82% of the blind was of 50 years and older [1-3]. The prevalence of blindness in Nepal was 0.84% in 1981 and 0.35% in 2012 whereas the prevalence of blindness in sudurpaschim province was higher than national average at 1.02% in 1981 and 0.41% in 2012. [4,5]

Cataract is the most common cause of blindness in the developing world [6–12], while retinal disorders are the most common cause of blindness in the developed world [14–20]. The leading cause of blindness in Nepal was cataract (62.2%) and in Sudurpaschim province it was (62.5%) followed by posterior segment disease and glaucoma. [5] Cataract was also the most common cause of blindness, followed by trachoma related blindness, according to the Nepal Blindness Survey conducted in 1981. A subsequent Rapid Assessment of Avoidable Blindness (RAAB) survey completed in 2012 determined cataract and posterior segment diseases as the leading cause of the blindness. Although cataracts have been repeatedly reported as the leading cause of blindness, posterior segment diseases and glaucoma were seen as more significant causes of blindness than trachoma related causes in the second survey. Strategic plans had been implemented to combat cataract and trachoma in the national and regional levels for a long period. Some other eye diseases related to systemic conditions like diabetes, age and occupational hazards have not been equally prioritized. Hence, there is a strong case to expect some shifts in the common causes of blindness and visual impairment in the region since the last survey was done about a decade back.

As the prevalence of blindness and visual impairment has not been assessed in Sudurpaschim province after 2012, we planned this study to determine prevalence of blindness and visual impairment among 50 years of age and older populations in this province in the recent scenario. This data will provide a measurement of the impact of eye care services in Sudurpaschim province in terms of changes in the prevalence of blindness, severe visual impairment (SVI), moderate visual impairment (MVI), and also help the government and stakeholders to develop focused strategies to combat specific causes of blindness as identified in the study.

## Materials and Methods

The study was conducted after ethical approval from Nepal Health Research Council (NHRC). This is a cross sectional population based survey using RAAB survey methodology [13]. The sample size was calculated for this study using the RAAB+DR V.6 software using the following indicators: current population 50 years and older 250,982 inhabitants, the assumed prevalence of blindness among this group as 2.9% with 20% tolerable error, a 1.4 cluster design effect for cluster of 35 people, 95% confidence intervals and 10% non-response rate. The total required sample size was estimated to be 4619, divided into 132 clusters each consisting 35 people completed age 50 years.

In this survey, we used multistage cluster sampling methodology. According to 2011 census data, approximately 15% of the population was 50 or older. Hence, we created a population unit of approximately 250 people to meet 35 study participants in one cluster. In the previous political division of Nepal, the village development committee (VDC) ward was the smallest population unit close to our desired cluster size. Therefore, all 3517 VDC wards in the survey area were listed, which served as the sampling frame for this study. In the first stage 132 population units were randomly selected as study clusters from the sampling frame.

In the second stage 35 eligible people in each cluster were randomly selected via compact segment sampling. The enumeration area was divided into segments of approximately equal population size with well-demarcated boundaries (using the map) so that each segment includes the desired cluster size of 35 people aged 50 or older, and one segment was randomly selected. In the selected study cluster, all eligible households were enrolled with door-to-door enumeration, until the desired number of people aged 50 and older were identified. If there were fewer than the required number of people aged 50 or older in the segment, an adjacent segment was randomly selected, and enrolment with data collection were continued until a total of 35 survey participants were enrolled.

An ophthalmologist-led team comprised of one ophthalmic assistant and one eye worker went to selected study clusters and examined this sample population. Ophthalmic assistant measured the visual acuity at a distance of 6 meters using tumbling-E opto types (Precision Vision, Villa Park, Illinois) with and without pinhole. People wearing glasses were tested with them and their visual acuity considered as presenting visual acuity. Anterior segment eye examination was carried out with torch light, and a portable slit lamp and fundus examination was done using a direct ophthalmoscope. If any abnormality requiring further examination investigation or treatment was detected, those patients were sent to Geta Eye Hospital or its nearest eye center for further management.

Blindness was defined as presenting visual acuity (PVA) in the better eye of less than 3/60, severe visual impairment (SVI) as PVA of less than 6/60 to 3/60, moderate visual impairment (MVI) as PVA of less than 6/18 to 6/60, and early visual impairment (EVI) as PVA less than 6/12 to 6/18, as per WHO criteria [21].

## RESULT

**Table 1:** Total participants in the study.

|  | Enrolled | Examined | Response rate<br>% |
| --- | --- | --- | --- |
| MALE | 2007 | 1995 | 99.4% |
| FEMALE | 2608 | 2578 | 98.8% |
| TOTAL | 4615 | 4573 | 99.1% |

A total of 4615 people aged 50 years or older were enrolled. Out of them 4573 (99.1%) underwent eye examination and data collection. Out of 4573, males were 1995 (43.63%) and females were 2578 (56.37%). Among the examined, 2214 (48.4%) were between the age of 50-59 years, 1233 (27%) were between the age of 60-69 years, 842 (18.4%) were between the age of 70-79 years and 284 (6.2%) were of 80 or older. These age groups that participated in the study were adjusted with census data from the same age group and presented as age and sex adjusted results.

**Table 2:** Age and Sex adjusted Prevalence of Blindness, SVI, MVI and EVI by bilateral presenting visual acuity.

|  | Male |  | Female |  | All |  |
| --- | --- | --- | --- | --- | --- | --- |
|  | n | %(95%CI) | n | %(95%CI) | n | %(95%CI) |
| Blindness PVA <3/60 | 639 | 0.5(0.2-0.9) | 857 | 0.7(0.4-1.0) | 1496 | 0.6(0.4-0.8) |
| SVI PVA (<6/60-3/60) | 1,298 | 1.0(0.5-1.5) | 1084 | 0.9(0.5-1.3) | 2382 | 1.0(0.6-1.3) |
| MVI PVA(<6/18-6/60) | 6,224 | 4.9(3.9-6.0) | 7183 | 5.7(4.5-7.0) | 13407 | 5.3(4.5-6.2) |
| EVI PVA(<6/12-6/18) | 8,967 | 7.1(5.8-8.4) | 8620 | 6.9(5.7-8.1) | 17602 | 7.0(6.0-8.0) |

The age and sex adjusted prevalence of blindness was 0.6%. The age and sex adjusted prevalence of severe visual impairment (SVI), moderate visual impairment (MVI) and early visual impairment (EVI) were 1.0%, 5.3% and 7.0% respectively.

**Table 3:**
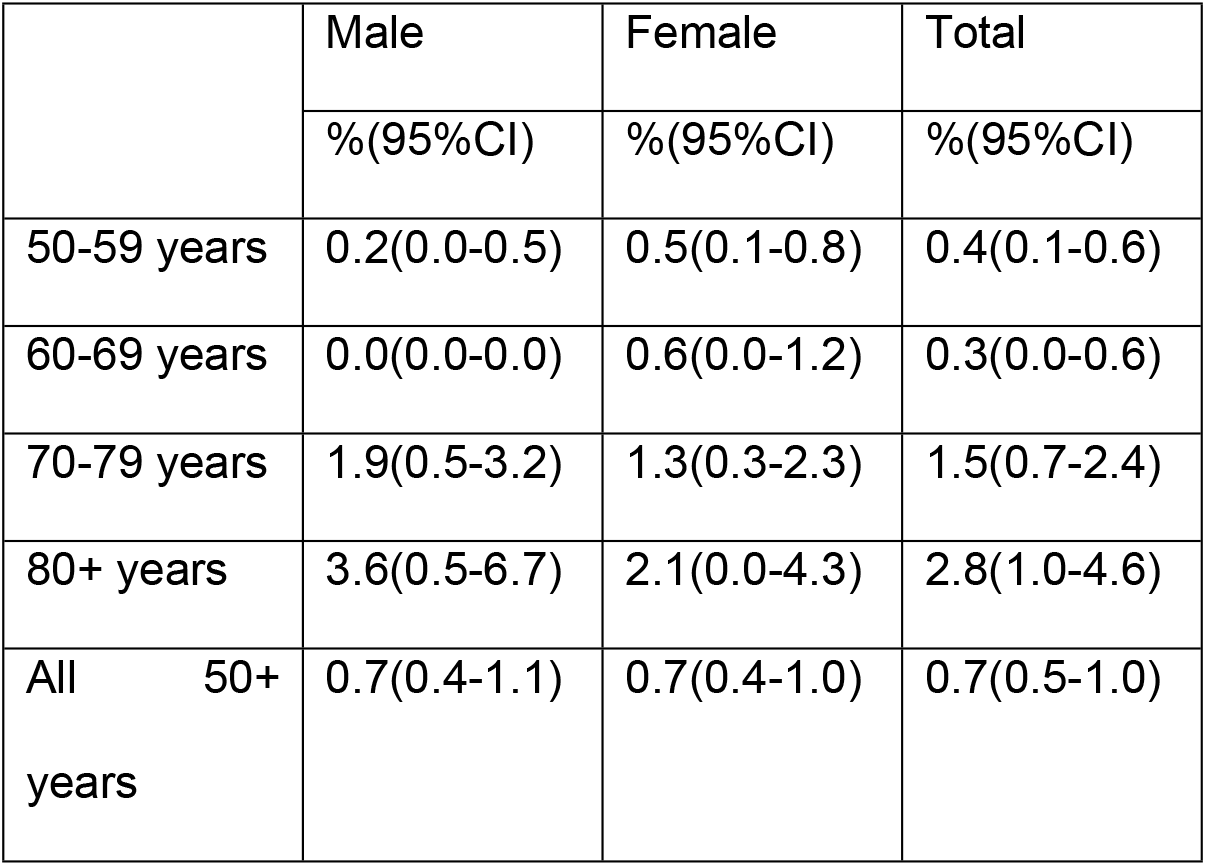
Crude Prevalence of blindness (PVA<3/60) by age group.

The prevalence of blindness was higher in the elderly population, with the highest prevalence among the age group 80+ years (2.8%, 95% CI 1.0-4.6) followed by age group 70-79 years (1.5%, 95% CI 0.7-2.4)

**Table 4:** Causes of blindness, SVI, MVI, EVI.

|  | **Blindness**<br>**(PVA<3/60)** |  | **SVI (PVA**<br>**<6/60-3/60)** |  | **MVI (PVA <6/18-**<br>**6/60)** |  | **EVI (PVA**<br>**<6/18-6/12)** |  |
| --- | --- | --- | --- | --- | --- | --- | --- | --- |
|  | n | % | n | % | n | % | n | % |
| Cataract untreated | 12 | 36.4% | 42 | 85.7% | 150 | 55.1% | 55 | 16.4% |
| Non trachomatous<br>corneal opacity | 8 | 24.2% | 0 | 0.0% | 6 | 2.2% | 6 | 1.8% |
| Glaucoma | 7 | 21.2% | 1 | 2.0% | 2 | 0.7% | 4 | 1.2% |
| All other globe/<br>CNS abnormalities | 3 | 9.1% | 0 | 0.0% | 0 | 0.0% | 0 | 0.0% |
| Age related<br>macular<br>degeneration | 2 | 6.1% | 2 | 4.1% | 12 | 4.4% | 2 | 0.6% |
| Other posterior<br>segment disease | 1 | 3.0% | 1 | 2.0% | 1 | 0.4% | 3 | 0.9% |
| Refractive error | 0 | 0.0% | 1 | 2.0% | 87 | 32% | 256 | 76.2% |
| Cataract surgical<br>complication | 0 | 0.0% | 1 | 2.0% | 11 | 4.0% | 8 | 2.4% |
| Aphakia<br>uncorrected | 0 | 0.0% | 1 | 2.0% | 2 | 0.7% | 1 | 0.3% |
| Trachomatous<br>corneal opacity | 0 | 0.0% | 0 | 0.0% | 1 | 0.4% | 1 | 0.3% |

Untreated cataract was by far the most common cause of blindness (36.4%), SVI (85.7%), MVI (55.1%) and EVI (16.4%) in our study. The other common causes of blindness after untreated cataract were non trachomatous corneal opacity (24.2%), glaucoma (21.2%), all other globe or central nervous system abnormalities (CNS) (9.1%), Age related macular degeneration (ARMD) (6.1%) and other posterior segment disease (3.0%).

## DISCUSSION

According to the census 2011, the Sudurpaschim Province of Nepal, which consists of nine districts, had a population of 2,543,349 people. The only tertiary level eye hospital in this province is Geta Eye Hospital (GEH). The hospital also runs one secondary level hospital, four surgical eye centers, and sixteen primary eye care centers in different parts of this province. This hospital has been providing eye care services through these centers and also through screening and surgical eye camps since its establishment in 1981. Apart from Geta Eye Hospital and its branches, the province has only one newly opened private eye hospital in the city area. Government hospitals and health centers have no provision for eye care services in the province. Hence, the bulk of eye care services in this province are provided by GEH and its affiliated hospital and eye centers. Hence this study was conducted to understand the impacts of eye care services in the province and also to understand major causes of blindness and visual impairment in the province to streamline future eye care strategies.

The total number of people aged 50 years or above enrolled in the study was 4615, and the total number of people examined was 4573, with a response rate of 99.1%. In a similar study done in Maldives, the response rate was 97.4% which is slightly lower than our study [23]. In another similar study done in west Nigeria, the response rate was 85% [24].

In our study, the age-sex adjusted prevalence of blindness among the people aged 50 years or above was 0.6% (95% CI, 0.4%-0.8%). Previous RAAB survey conducted in this province in 2012, reported the age-sex adjusted prevalence of blindness of 2.7% (95% CI, 1.8%-3.7%). One of the major reasons for this reduction in blindness is the successful implication of a community based approach to eye care with special focus on outreach surgical camps, which have been able to provide good outcomes [25]. The prevalence of SVI 1.0% (95% CI, 0.6-1.3%) and MVI 5.3%, (95% CI, 4.5-6.2%) have also decreased from previous rates of 2% and 7.6% respectively [5]. The prevalence rate of blindness and visual impairments noted in our study also compares favorably to those reported from some of the developing countries in the region. The age–sex adjusted prevalence of blindness, severe visual impairment and moderate visual impairment in Malaysia were 1.2% (95% CI, 1.0% - 1.4%), 1.0% (95% CI, 0.8% - 1.2%) and 5.9% (95% CI, 5.3% -6.5%) respectively [26]. In a study conducted in Bihar, India, the age – sex adjusted prevalence of blindness among people 50 years or above was 2.2% (95% CI, 1.6-2.8). The prevalence of SVI, MVI, and EVI were 3.4% (95% CI, 2.6-4.3%), 18.3% (95% CI, 16.8-19.9%) and 16.9% (95% CI, 15.1-18.7) respectively [27]

The prevalence of blindness was higher in females 0.7% (95% CI, 0.4%-1.0%) compared to males 0.5% (95% CI, 0.2%-0.9%) which may be due to gender based differences in eye health and service accessibility. The prevalence of blindness in this study was highest in those aged 80 or older (2.8%) followed by 1.5% in those aged 70-79 years. In a similar study done in Maldives in 2018, it was 13.1%, and 4.2%, in a population of aged 80 or older and 70-79 years respectively [23]. Both studies show a higher prevalence of blindness in older age group.

The proportion of cataract related blindness has decreased from 62.5 % in 2012 [5] to 36.4% in our study. One of the important reason for this decrease in the prevalence of cataract related blindness is the adoption of high volume cataract surgery protocols by Geta Eye Hospital to be able to address the large backlog as evidenced in the previous study. These protocols have also been able to provide high quality outcomes, helping to decrease the prevalence of blindness and visual improvement in the province. [25, 28]

Despite this progress, cataract is still the main cause of blindness (36.4%) followed by non trachomatous corneal opacity (24.2%), glaucoma (21.2%), all other globe or CNS abnormality (9.1%), ARMD (6.1%), other posterior segment disease (3%). Again, cataract is the main cause in SVI (85.7%) and MVI (55.1%) category. This shows that cataract intervention programs still need to be an important part of the eye care services, along with efforts to tackle other rising causes of blindness and visual impairment in this province. Cataract was the major cause of SVI in India (77.5%), Bangladesh (73.6%) and Bhutan (74.1%) similar to our study [29]. According to the same report, cataract was the main cause of MVI in India (58.1%) and Bhutan (57.1%), whereas in Bangladesh uncorrected refractive error (63.6%) was the main cause for MVI [29].

The second major cause of blindness in this study is non-trachomatous scar (24.2%), which was only 2.8% in previous study [5]. Similarly, glaucoma has come up as the third major cause of blindness in this study, which has increased to 21.2% from 9.7% in the previous study [5]. These findings show that vision loss due to other eye diseases has been increasing in proportion as we have been able to reduce cataract related blindness to some extent in the community. Hence, we also need to develop strategic plans to combat these eye problems in the future as we did for cataracts and trachoma. There is not a single case of trachomatous scar in blindness category in this study, whereas there were 4.2% in the previous study. Nepal has already been declared as a trachoma eliminated country, thanks to the efforts of both government and various non-governmental organizations working together to eliminate trachoma.

### Limitations

Due to Corona virus disease of 2019 pandemic, the survey had been interrupted for about 6 months and completed with precautions later on.

## Conclusion

The prevalence of blindness was significantly reduced in the province in comparison to the previous survey. Although cataract prevalence has decreased, it still remains the major cause of blindness, SVI and MVI followed by corneal opacities and glaucoma, which are emerging as second and third major causes of blindness, respectively. Hence, higher priority should be required on cataract, corneal blindness and glaucoma in future interventions to eliminate blindness in sudurpaschim province of Nepal.

## Data Availability

All relevant data are within the manuscript and its Supporting Information files.

## ACKNOWLEDGEMENTS

We wish to thank Mr Ram Prasad Kandel, Country Director of SEVA Nepal for coordination with SEVA Foundation USA for financial support for the study, Ophthalmologists of Geta Eye Hospital Dr Smadh Adhikari, Dr Sebanta Shrestha, Dr Sunil Thapa and all staffs participated in data collection for the study. We would like to thank to all subjects who voluntarily participated in the study.

